# The penetrance of age-related monogenic disease depends on ascertainment context

**DOI:** 10.1101/2021.06.28.21259641

**Authors:** Uyenlinh L Mirshahi, Kevin Colclough, Caroline F Wright, Andrew R Wood, Robin N Beaumont, Jessica Tyrrell, Thomas W Laver, Richard Stahl, Alicia Golden, Jessica M Goehringer, on behalf of Geisinger-Regeneron DiscovEHR Collaboration, Timothy F Frayling, Andrew T Hattersley, David J Carey, Michael N Weedon, Kashyap A Patel

## Abstract

**BACKGROUND:** Accurate penetrance of monogenic disorders is often unknown due to a phenotype-first approach to genetic testing. Here, we use a genotype-first approach in four large cohorts with different ascertainment contexts to accurately estimate penetrance of the three commonest causes of monogenic diabetes, Maturity Onset Diabetes of the Young (MODY). We contrast *HNF1A-*MODY / *HNF4A*-MODY which causes an age-related progressive diabetes and *GCK-*MODY, which causes life-long mild hyperglycaemia.

**METHODS:** We analysed clinical and genetic sequencing data from four different cohorts: 1742 probands referred for clinical MODY testing; 2194 family members of the MODY probands; 132,194 individuals from an American hospital-based cohort; and 198,748 individuals from a UK population-based cohort.

**RESULTS:** Age-related penetrance of diabetes for pathogenic variants in *HNF1A* and *HNF4A* was substantially lower in the clinically unselected cohorts compared to clinically referred probands (ranging from 32% to 98% at age 40yrs for *HNF1A*, and 21% to 99% for *HNF4A*). The background rate of diabetes, but not clinical features or variant type, explained the reduced penetrance in the unselected cohorts. In contrast, penetrance of mild hyperglycaemia for pathogenic *GCK* variants was similarly high across cohorts (ranging from 89 to 97%) despite substantial variation in the background rates of diabetes.

**CONCLUSIONS:** Ascertainment context is crucial when interpreting the consequences of monogenic variants for age-related variably penetrant disorders. This finding has important implications for opportunistic screening during genomic testing.

## Introduction

Interpreting the medical consequences of rare variants in genes that cause monogenic disorders is extremely challenging. One of the major issues is that the penetrance – the absolute risk of developing the disease when an individual carries a pathogenic variant – is often not accurately known. Until recently there has been a phenotype-first approach to sequencing monogenic disease genes. These genes are identified by studying highly selected, small cohorts of patients and families with a specific condition^1-5^. Diagnostic testing of these genes is then usually only performed on individuals suspected to have the genetic disorder^6,7^. This testing strategy leads to the conclusion that the conditions are highly penetrant (i.e., most individuals with the variant will get the condition)^1-5^. Accurate estimates of penetrance are vital for testing and counselling family members and are becoming increasingly important as exome/genome sequencing becomes ubiquitous and more individuals are identified with pathogenic variants before they develop the corresponding phenotype.

The recent availability of large-scale exome sequencing datasets from population-based cohorts allows a genotype-first approach to estimating penetrance. Large-scale research studies such as the UK Biobank (N=500,000) and health centre studies such as the Geisinger/DiscovEHR (N=180,000) have extensive clinical and health record data^8-10^. The increasing availability of exome sequencing data in these studies allows the assessment of penetrance of monogenic disease variants in different settings and with different ascertainment criteria.

Maturity Onset Diabetes of the Young (MODY) provides an excellent disease model to assess the role of ascertainment on penetrance estimates. MODY is an autosomal dominant form of monogenic disease, classically described as a non-insulin dependent diabetes that is usually diagnosed before 25 years of age^11^. It is not a lethal disease, so patients with the phenotype will appear in older population cohorts. The phenotype of diabetes and the biomarkers used to define diabetes, such as blood glucose and HbA1c, are often well captured in population cohorts^8,9,12^. There is a worldwide agreed definition of diabetes based on these biomarkers, which means that the diabetes status of all the participants is reliably known. Pathogenic variants in the genes that most commonly cause MODY (*HNF1A, HNF4A* and *GCK*), which account for >90% of all cases, are extremely well defined^13-15^. Additionally, MODY has distinct subtypes which provide an opportunity to contrast age-related progressive disease (*HNF1A/4A*-MODY) with a lifelong stable disease (*GCK*-MODY). These reasons along with the existence of a large cohort of clinically referred cases for comparison make MODY an excellent example for assessing the role of ascertainment on monogenic disease.

In this study, we use MODY as an example to comprehensively assess the penetrance of rare genetic variants in four different settings and with different ascertainment contexts. We used a clinically selected cohort (MODY proband cohort) and their family members (family member cohort), a heath-system-based cohort (Geisinger cohort), and a population-based cohort (UK Biobank) to assess the penetrance of diabetes for the three most common causes of MODY.

## Methods

### Study populations

#### MODY probands cohort

We included 1742 probands up to age 85 who were referred for genetic testing at the Molecular Genomics Laboratory at the Royal Devon and Exeter Hospital, Exeter, UK with a clinical suspicion of MODY from routine primary or specialist clinical care from the UK. They were subsequently found to harbour a pathogenic variant in *HNF1A* (N=661), *HNF4A* (N=142) or *GCK* (N=939). Informed consent was obtained from the probands or their parents/guardians and the study was approved by the North Wales ethics committee. The clinical features of these individuals at referral for genetic testing are shown in Table S1 in the Supplementary Appendix.

#### Family members cohort

The family member cohort comprises the individuals up to age 85 who were related (up to third degree) to the MODY probands (N=2194). These individuals were referred from routine clinical care for family genetic screening to the Molecular Genetics Laboratory at the Royal Devon and Exeter Hospital. This included family members for *HNF1A*-MODY probands (N=954), *HNF4A*-MODY probands (N=253) and *GCK*-MODY probands (N=987). The clinical features of these individuals are shown in Table S2.

#### Geisinger cohort

The Geisinger cohort is a heath-system-based cohort from the USA consisted of 132,194 individuals up to age 85 years who sought healthcare at an outpatient and/or inpatient facility within Geisinger, a health care provider to central and north-eastern Pennsylvania, USA. Individuals consented to participate in the MyCode Community Initiative to create a biorepository of blood, serum and DNA samples for broad research use, including genomic analysis^9^. MyCode samples are linked to Geisinger electronic health records (EHR). We used the routinely collected data including clinical diagnosis, procedures, medications, and laboratory results from MyCode participants during their encounters with Geisinger providers in this study. Individuals where age of diabetes diagnosis could not be determined from their EHR were excluded from the study (N=2171/33,415 of all diabetes cases). Genetic analysis was carried out as part of the DiscovEHR collaboration between Geisinger and the Regeneron Genetics Centre by microarray genotyping and exome sequencing. This study was reviewed by the Geisinger Institutional Review Board and determined as not including human subject research as defined in 45CFR46.102(f) in written consent (Study #2016-0269). Of the total cohort, 87,225 (66%) were unrelated up to third-degree relationship. The cohort characteristics at recruitment for exome analysis are summarised in Table S1 and have been described extensively^10,16^.

#### UK Biobank

UK Biobank is a population-based cohort from the UK with deep phenotyping data and genetic data for around 500,000 individuals aged 40-70 years at recruitment^12^. Participants provided a range of information via questionnaires and interviews including diabetes status. Additionally, a panel of biomarkers was measured from blood and urine, including random blood glucose and HbA1c^8^. Phenotypes were derived from medical history interviews, in- and outpatient ICD9 and ICD10 codes, operation codes, and death registry data. A subset of ∼200,000 DNA samples from UK Biobank participants underwent exome sequencing; this dataset was recently made available for research^17^. The UK Biobank resource was approved by the UK Biobank Research Ethics Committee and all participants provided written informed consent to participate. Individuals with missing age at diabetes diagnosis were excluded (N=1135/12,569 of all diabetes cases). Table S1 described the clinical characteristics at recruitment for 198,748 individuals included in the study, of whom 184,142 (93%) were unrelated up to third-degree relationship.

### Definition of diabetes and mild hyperglycaemia

Diabetes was defined as being one or more of self-reported by participants or having an ICD9/10 code for diabetes or being on a diabetes treatment or having HbA1c ≥48 mmol/mol before recruitment^18,19^. Mild hyperglycaemia was defined accordingly to American Diabetes Association definition of prediabetes (HbA1c ≥39 mmol/mol or a fasting glucose ≥5.6 mmol/L)^18^.

### Age at diagnosis of diabetes

Age of diabetes diagnosis was self-reported in the UK Biobank at recruitment. For the Geisinger cohort, we used age at first evidence of diabetes diagnosis from ICD9/10 code or start of anti-diabetic medication or first HbA1c measurement >48 mmol/mol.

### Genetic analysis

#### MODY probands and family members

Sanger sequencing or targeted next-generation sequencing was used to undertake genetic analysis for these cases. The detailed method for these assays has been described previously^6^. Variants in *HNF1A, HNF4A* and *GCK* were analysed by the clinical scientists at the Molecular Genetics Laboratory at the Royal Devon and Exeter Hospital as part of the routine diagnostic care. All probands included in this study had variants classified as likely pathogenic (class 4) or pathogenic (class 5). Interpretation and classification of sequence variants were undertaken based on the American College of Medical Genetics and Genomics (ACMG)/Association of Molecular Pathology (AMP) guidelines^20^. The list of the included variants is shown in Supplementary table 3. We annotated variants by clinically used transcripts (NM_000545.5 for *HNF1A*, NM_175914.4 for *HNF4A* and NM_000162.3 for *GCK)*.

#### Geisinger cohort

Individuals underwent exome sequencing as part of the DiscovEHR collaboration of Geisinger (Danville, PA) and the Regeneron Genetics Centre (Tarrytown, NY)^10^. The detailed method for exome sequencing has been described previously^21^. Quality controls included filtering for samples of variants with read depth ≥ 10 (insertions and/or deletions, indels) or ≥ 7 (single nucleotide variants, SNVs), alternate allele balance >15% for SNVs or >20% for indels, and alternate allele reads >5.

#### UK Biobank

Detailed sequencing methodology for UK Biobank samples is provided by Szustakowski *et al*.^17^ and is available at https://biobank.ctsu.ox.ac.uk/showcase/label.cgi?id=170. Briefly, exomes were captured with the IDT xGen Exome Research Panel v1.0 which targeted 39Mbp of the human genome with coverage exceeds on average 20x on 95.6% of sites. The OQFE protocol was used for mapping and variant calling to the GRCh38 reference. We included variants that had individual and variant missingness <10%, Hardy Weinberg Equilibrium p-value >10^−15^, minimum read depth of 7 for SNVs and 10 for indels, and at least one sample per site passed the allele balance threshold > 15% for SNVs and 20% for indels.

#### Variant annotation and classification in UK Biobank and Geisinger cohorts

Variants were annotated by AlaMut batch software v1.8 (Interactive Biosoftware, France) using clinical transcripts for *HNF1A, HNF4A* and *GCK* as listed above. *HNF1A, HNF4A* and *GCK*-MODY is caused by haploinsufficiency in these genes^13,14^ and heterozygous pathogenic variants can be missense or protein-truncating^13,14^. We defined a protein-truncating variant (PTV) as a variant that is predicted to cause a premature stop gain, a frameshift, or abolish a canonical splice site (−2 or +2 bp from exon boundary). In this study, we excluded PTVs in the last exon of each gene and only those deemed to be high confidence by the Loss-Of-Function Transcript Effect Estimator (LOFTEE)^22^ were retained.

We reviewed all heterozygous missense in UK Biobank and the Geisinger cohort that were observed at minor allele frequency (MAF) <0.001 in gnomAD v2 (N=141,456)^22^ and in each study cohort respectively. All PTVs irrespective of the MAF were reviewed in UK biobank and the Geisinger cohort. ACMG/AMP guidelines were used to classify variants as pathogenic/likely pathogenic^20^ as follows: all PTVs were classified as pathogenic; missense variants were classified as pathogenic if the variant is seen in a MODY proband and was classified as pathogenic or likely pathogenic based on ACMG/AMP guideline by clinical scientists at Exeter Molecular Genetic laboratory as part of routine clinical diagnostic care and was very rare in the population (maximum allele count of 2 in gnomADv2, MAF <1.4×10^−5^). Missense variants that were not seen in MODY probands were classified as either VUS or benign accordingly to ACMG/AMP guideline.

Three researchers independently manually reviewed sequence read data for all the pathogenic variants (missense and PTVs) in Integrative Genomics Viewer (IGV)^23^ to remove false positive variants. The variants considered to be excellent quality by all three researchers were included in the analysis.

#### Sanger sequencing validation of the HNF1A c-insertion variant in the Geisinger cohort

The most common *HNF1A* pathogenic variant is a frameshift variant (p.Pro289AlafsTer28) in exon 4 due to an insertion of a C. This variant is difficult to detect robustly in exome/genome sequencing data due to the location in a repetitive poly-C tract and the presence of a common variant at the end of the tract (rs56348580 G>C, MAF=0.26). There were 23 frameshift variants detected in this region in the UK Biobank and 29 in the Geisinger cohort. We Sanger sequenced all individuals that carried a putative c-insertion variant in the Geisinger cohort and only 4 (17%) were confirmed. We therefore excluded all *HNF1A* c-insertion variants from the UK Biobank cohort as we were unable to perform Sanger sequencing confirmation.

### Statistical analysis

Kaplan-Meier survival estimate was used to compute the age-dependent penetrance of diabetes in *HNF1A* and *HNF4A* carriers. Log-rank test for equality was used to compare the penetrance of diabetes between the groups. Cox’s regression was used to compute hazard ratio for developing diabetes with or without adjustment of covariates. Fisher’s exact test was used to compare the penetrance of mild hyperglycaemia in *GCK* carriers between the cohorts. We used linear regression to compare fasting glucose and HbA1c levels with and without adjustment of covariates between the *GCK* carriers from different cohorts. We used Cochran’s Q test to assess heterogeneity between the study cohorts. All the analysis was performed using Stata 16 (College Station, Texas, USA).

## Results

### Penetrance of pathogenic *HNF1A* variants is lower in clinically unselected cohorts compared to a clinically ascertained cohort

We assessed the age-related penetrance of pathogenic *HNF1A* variants in MODY probands (N=661), their family members (N=954), a health-system-based cohort (Geisinger cohort N=132,194) and a population-based cohort (UK Biobank N=198,748). The different background rate of diabetes highlights the different ascertainment of these cohorts (100%, 61%, 24% and 6%, respectively). We identified 661, 622, 14, 22 people with a pathogenic variant in *HNF1A* in these cohorts, respectively (Tables S3 and S4).

Kaplan-Meier analysis demonstrated the penetrance of diabetes for pathogenic *HNF1A* variants was lower in the family members, the Geisinger cohort and UK Biobank compared to the *HNF1A*-MODY probands (log-rank test, all *P*<3×10^−9^; Fig. 1A, Fig. S1). For example, by age 40 years, 98% (95%CI 97-99%) of probands, 86% (83-89%) of family members, 49% (25-78%) of the Geisinger cohort and 32% (17-55%) of UK Biobank carriers were diagnosed with diabetes respectively (Fig. 1B). The penetrance remained lower in these cohorts compared to probands after the adjustment of age, Body Mass Index (BMI), sex, parental diabetes, and variant type (PTV vs missense) in a multivariable Cox proportional hazard model (Table S5). The results were also similar when the analysis was restricted to unrelated individuals of European ancestry (Table S5).

**Figure 1:**
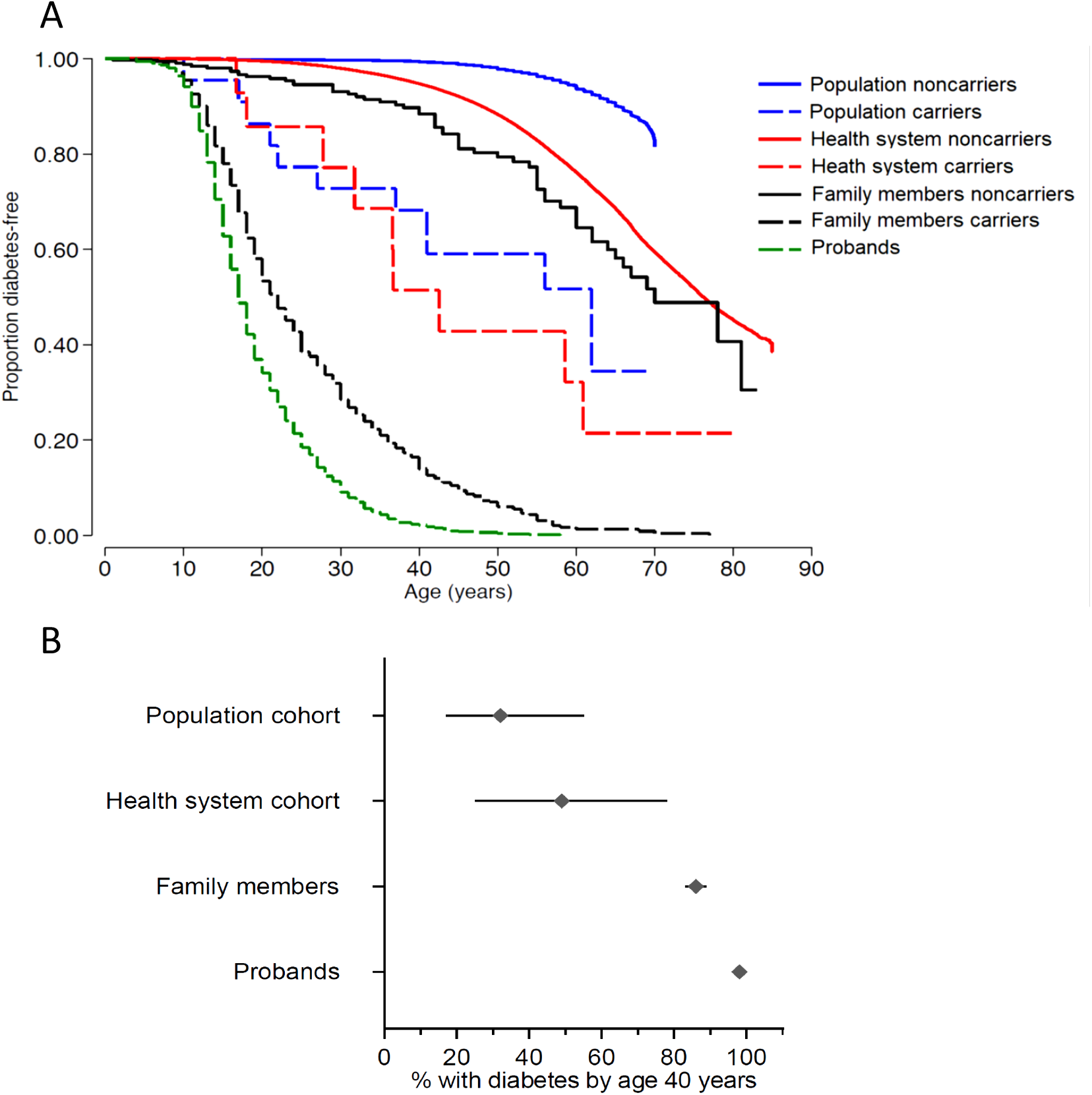
Penetrance of pathogenic *HNF1A* variants is lower in clinically unselected cohorts compared to a clinically ascertained cohort. A) Kaplan Meier survival curves of diabetes for *HNF1A* variant carriers (dashed line) and noncarriers (solid line) in four study cohorts. Analysis included probands (N=661), their family members carriers (N=954) and noncarriers (N=332), carriers (N=14) and noncarriers (N=132,148) of from Geisinger health system cohort and carriers (N=22) and noncarriers (N=198,726) from UK Biobank population cohort. The log rank test p value for penetrance of diabetes for probands versus family member, Geisinger cohort and UK Biobank *HNF1A* carriers was 3×10^−26^, 3×10^−09^, 5×10^−16^, respectively. B) Penetrance of diabetes for pathogenic *HNF1A* variant carriers in all four cohorts at age 40 years with 95% CI.

### Penetrance of pathogenic *HNF4A* variants is lower in clinically unselected cohorts compared to a clinically ascertained cohort

We next assessed the age-related penetrance of pathogenic *HNF4A* variants in the same four cohorts (MODY probands [N=142], their family members [N=253], Geisinger cohort and UK Biobank). We identified 142, 169, 20, 29 individuals with a pathogenic variant in *HNF4A*, respectively (Tables S3 and S6).

For individuals with a *HNF4* pathogenic variant, the age-related penetrance of diabetes was lower in the family members, the Geisinger cohort and the UK Biobank compared to MODY probands (log-rank test, all *P*<8×10^−11^; Fig. 2A, Fig. S2). For example, by age 50 years, 99% (95%CI 96-100%) of probands, 90% (83-95%) of family members, 26% (10-56%) of the Geisinger cohort and 21% (10-41%) of UK Biobank carriers developed diabetes respectively (Fig. 2B). The lower penetrance in clinically unselected cohorts was maintained after the adjustment of age, BMI, sex, parental diabetes, and variant type (PTV vs missense) in a multivariable Cox proportional hazard model (Table S7). The result was also similar when the analysis was restricted to unrelated individuals of European ancestry (Table S7).

**Figure 2:**
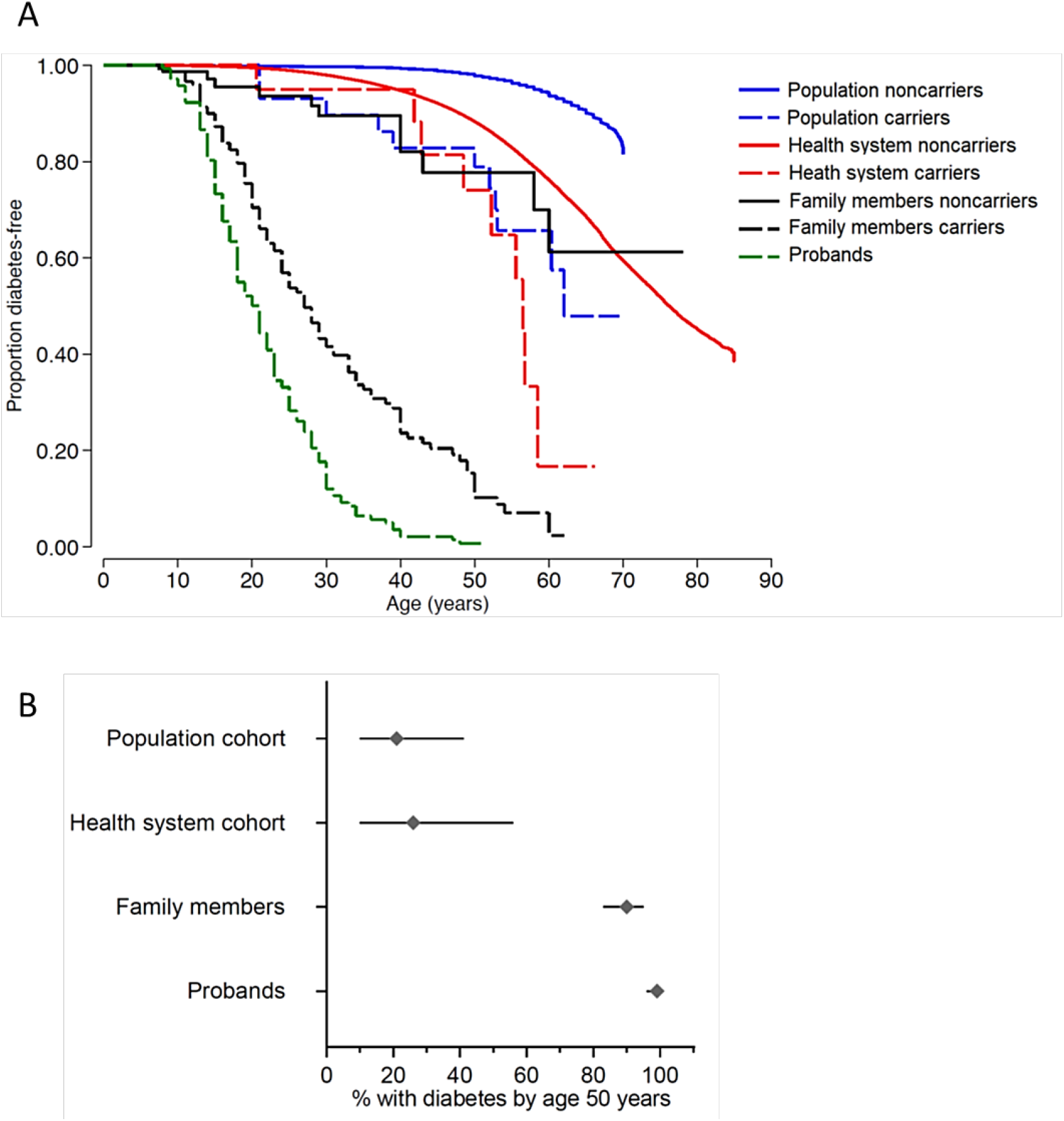
Penetrance of pathogenic *HNF4A* variants is lower in clinically unselected cohorts compared to a clinically ascertained cohort. A) Kaplan Meier survival curves of diabetes for *HNF4A* variant carriers (dashed line) and noncarriers (solid line) in four study cohorts. Analysis included probands (N=142), their family members carriers (N=253) and noncarriers (N=84), carriers (N=20) and noncarriers (N=132,174) from Geisinger health system cohort and carriers (N=29) and noncarriers (N=198,719) from UK Biobank population cohort. The log rank test p value for penetrance of diabetes for probands versus family member, Geisinger cohort and UK Biobank *HNF4A* carriers was 8×10^−11^, 2×10^−14^, 3×10^−19^, respectively. B) Penetrance of diabetes for pathogenic *HNF4A* variant carriers in all four cohorts at age 50 years with 95%CI.

### The reduced penetrance of diabetes in clinically unselected cohort is not due to differences in pathogenic variants among these cohorts

We conducted multiple sensitivity analysis to assess the impact of variant heterogeneity on our results. We first analysed only carriers with PTVs in all cohorts and limited the MODY probands to the variants that were seen in the comparison cohorts. Both these analyses showed that the clinically unselected cohort had lower penetrance for *HNF1A* and *HNF4A* pathogenic variants (log rank test, all *P*<1×10^−4^ vs. probands for PTVs, and *P*<2×10^−4^ vs. probands for missense for *HNF1A;* and all *P*<0.01 for PTVs and *P*<1×10^−3^ for missense for *HNF4A*; Figs. S3 and S4). To completely remove the possible differential effect of variants, we assessed penetrance by restricting the analysis to a single pathogenic variant across the cohort. We did not have enough carriers of a single pathogenic variant in *HNF1A* or *HNF4A* for this analysis. However, we had adequate carriers for the pathogenic but distinct *HNF4A* MODY subtype (including lower penetrance) caused by p.(R114W) variant^24^ (N=37, 43, 24 and 58 in MODY probands, proband family members, Geisinger, and UK Biobank exome sequenced cohort, respectively). Penetrance of diabetes in carriers of *HNF4A* p.(R114W) variant was lower in the unselected cohorts compared to MODY probands (log rank test, all *P*<1×10^−9^; Fig. S5 and Table S8). These data together suggest that the lower penetrance of diabetes in clinically unselected cohort is not due to different pathogenic variants in these cohorts.

### The difference in the prevalence of diabetes in the study cohorts explains the difference in the age-dependent penetrance in carriers of pathogenic *HNF1A/4A* variants

The standard definition of penetrance (“the absolute risk of developing a disease in individuals with a pathogenic variant”)^25^ does not take into account the context in which the variant carriers were identified. We hypothesised that the differences in the prevalence of diabetes in our study cohorts (ascertainment context) may explain the observed variation in the penetrance (absolute risk) of diabetes. To assess this, we calculated cox proportional hazard ratios (HR) for developing diabetes in *HNF1A/4A* carriers relative to non-carriers in each of the three clinically unselected cohort. HR for all three unselected cohort was broadly similar for *HNF1A* carriers albeit slightly lower in the Geisinger cohort (11 [95%CI 8-15] for family members, 4 [2-8] for Geisinger cohort, 16 [9-28] for UK Biobank) (Fig. 3A and Table S9). Similar results were seen for *HNF4A* carriers (8 [4-14] for family members, 4 [2-7] for Geisinger cohort, 8 [4-14] for UK biobank, Fig. 3B), and carriers with *HNF4A* p.(R114W) variant (1 [0.6-3], 3 [2-5], 9 [0.8-4] respectively, Fig. 3C and Table S9). We observed similar results when the analyses were limited to unrelated individuals of European ancestry (Table S9). These data together suggest that difference in ascertainment was the major reason for the reduced penetrance seen in the clinically unselected cohorts and can largely be explained by the different background rates of diabetes.

**Figure 3:**
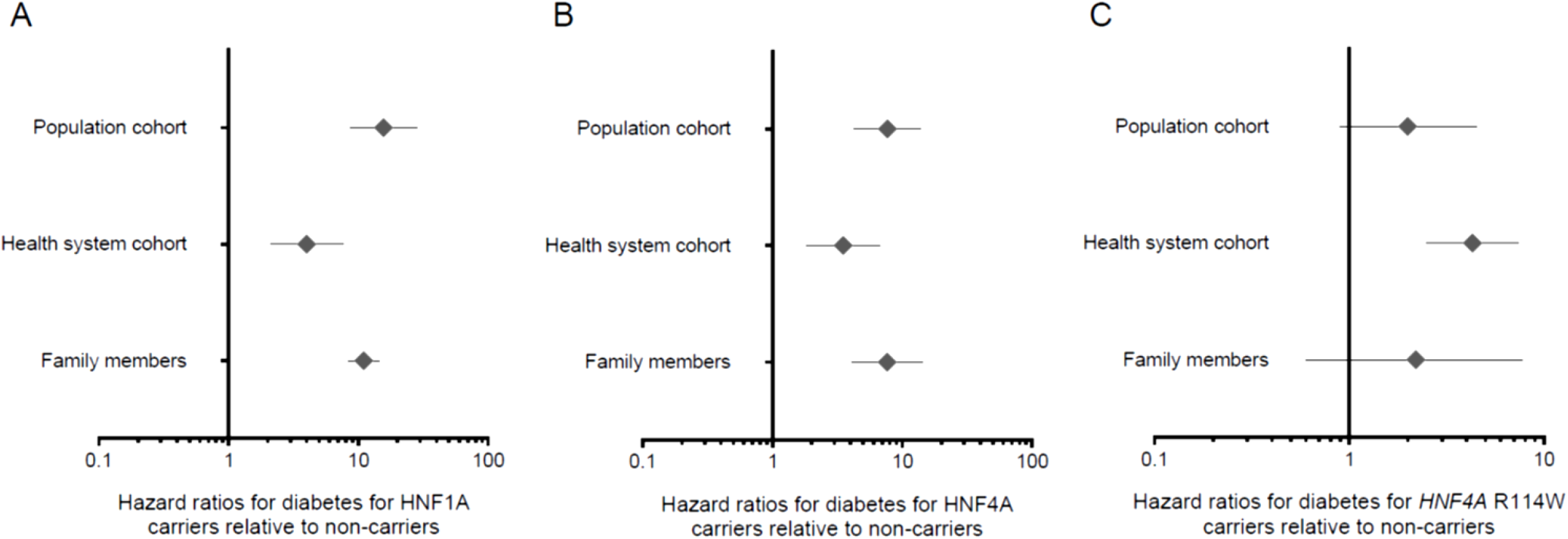
Cox proportional Hazard ratio for diabetes for pathogenic *HNF1A, HNF4A*, and *HNF4A* p.R114W variant carriers relative to noncarriers are largely similar in the clinically unselected cohorts. A) Graph showed Cox proportional Hazard ratio and 95%CI for diabetes for carriers of pathogenic *HNF1A* variants relative to noncarriers for family members, Geisinger health system cohort and UK Biobank population cohort. B) Graph showing the Cox proportional Hazard ratio and 95%CI for diabetes for carriers of pathogenic *HNF4A* variants for same three cohorts. C) Graph showing the Cox proportional Hazard ratio and 95%CI for diabetes for carriers of pathogenic *HNF4A* p.(R114W) pathogenic variant for same three cohorts.

### Penetrance of pathogenic *GCK* variants is not affected by ascertainment context

We next went on to assess the penetrance of pathogenic GCK variants as an example of life-long phenotype unlike the age-related phenotype of HNF1A/HNF4A-MODY. *GCK*-MODY causes life-long stable mild fasting hyperglycaemia from birth with a modest increase with age rather than true progressive diabetes as seen in *HNF1A/4A*-MODY^26-28^. Therefore, its penetrance is assessed by presence of mild hyperglycaemia (defined in this study as fasting blood glucose ≥5.6 mmol/L and/or HbA1c ≥39 mmol/mol)^18^.

We analysed penetrance of mild hyperglycaemia in the MODY probands, their family members, the Geisinger cohort and UK Biobank. We identified 939, 723, 32 and 83 carriers of a pathogenic variant in *GCK* in these cohorts, respectively (Tables S3 and 10).

The penetrance of mild hyperglycaemia was 97% (95%CI 96-98%) for *GCK*-MODY probands. The penetrance of *GCK* carriers in the unselected cohorts was similar to probands (family members 96% [94-98], *P*=0.30, Geisinger cohort 89% [71-98], *P*=0.04, UK Biobank 96% [90-99], *P*=0.48) despite the different prevalence of diabetes (46%, 24% and 6%) and mild hyperglycaemia (83%, 52% and 34% respectively) in the clinically unselected cohorts (Fig. 4A). Despite the lower and different background levels of HbA1c across the clinically unselected cohorts (mean HbA1c of 47.1, 44.9 and 38.2 mmol/L, respectively), the mean HbA1c of *GCK-MODY* probands was similar to those with GCK pathogenic variants from the Geisinger cohort (46.1 [95%CI 45.7-46.6] vs. 48.3 [44.3-52.3], *p*=0.1) and UK Biobank (47.5 [46.5-48.5], *p*=0.09) and marginally lower than that of family members (48.4 [47.5-49.3], *P*<0.0001 unadjusted, and *P*>0.05 for after adjustment of age, sex and BMI) (Fig. 4B; Table S11). Fasting blood glucose analysis also showed similar results as HbA1c (Fig. 4C, Table S12). Analyses restricting to unrelated individuals of European ancestry also showed equivalent results for HbA1c and fasting blood glucose (Tables S11, S12 and S13).

**Figure 4:**
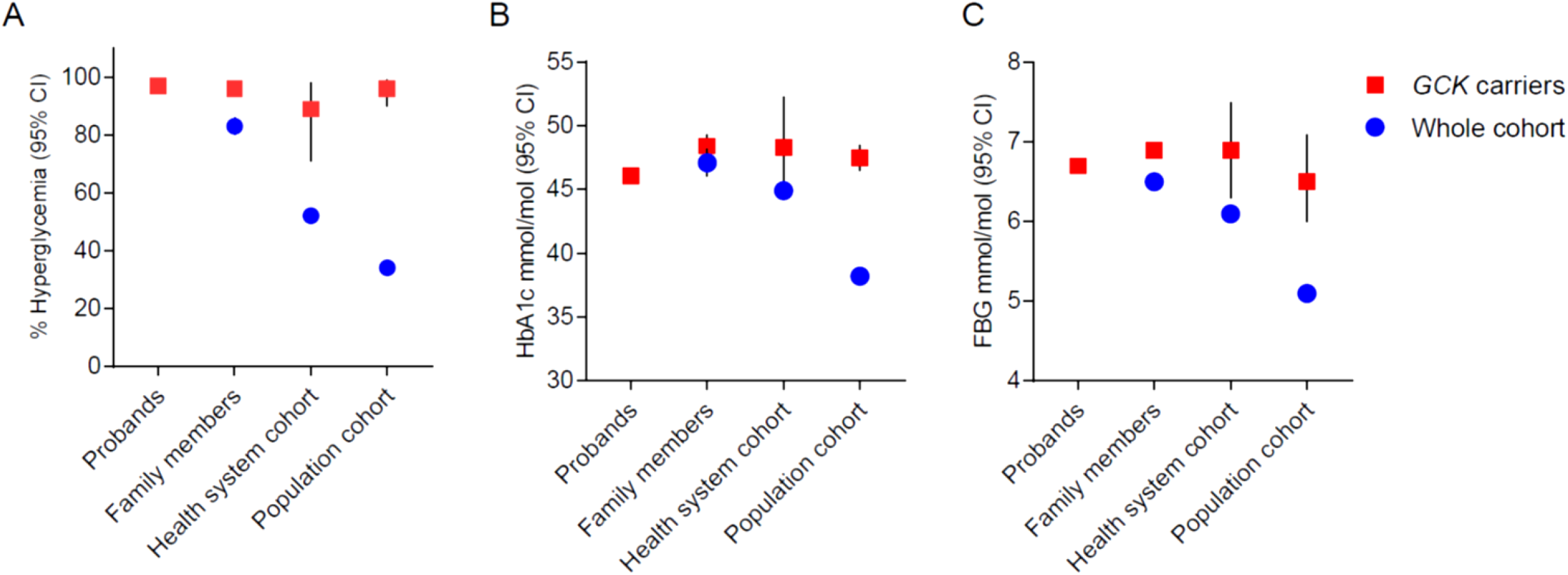
Penetrance of mild hyperglycaemia for pathogenic *GCK* variants is similar in clinically selected and unselected cohorts. A) Proportion and 95%CI of individuals with mild hyperglycaemia (HbA1c ≥39 mmol/mol and/or a fasting glucose ≥5.6 mmol/L, definition of prediabetes by American Diabetes Association) for probands of pathogenic *GCK* variants (N=939) and carriers of pathogenic *GCK* variants in their family members (N=987), Geisinger heath system cohort (N=32) and UK biobank population cohort (N=83) as red square. Background rate of mild hyperglycaemia and 95%CI in each unselected cohort is also shown as blue circle. B) Mean HbA1c and 95%CI for *GCK*-MODY probands and carriers of pathogenic *GCK* variants in their family members, Geisinger cohort and UK Biobank as red square. Cohort mean HbA1c and 95%CI for each unselected cohort is shown as blue circle C) Mean fasting blood glucose and 95%CI for *GCK*-MODY probands and carriers of pathogenic *GCK* variants in their family members, Geisinger cohort and UK Biobank as red square. Cohort mean fasting blood glucose and 95%CI for each unselected cohort is shown as blue circle.

## Discussion

Using four different cohorts with different ascertainment contexts, we have shown that age-related penetrance of two of the common causes of monogenic diabetes (*HNF1A*/*HNF4A*) is reduced in clinically unselected cohorts. This reduction in penetrance is consistent with the lower background rate of diabetes in these cohorts. In contrast, the age-independent penetrance of *GCK*-MODY was similar in all four cohorts irrespective of ascertainment context.

Pathogenic variants in *HNF1A* and *HNF4A* account for >66% of maturity-onset diabetes of the young. They are regarded as highly penetrant forms of diabetes with most individuals diagnosed before the age of 25 years^11,13^. Outside of our clinically referred cohort, we have shown that the penetrance is substantially lower than previously thought. Most individuals with *HNF1A* and *HNF4A*-MODY in two clinically unselected cohorts from the USA and UK did not develop diabetes even by 50 years of age (∼50% and ∼80% in Geisinger and UK Biobank, respectively). This suggests that phenotype-first studies of probands provide an upper limit of penetrance, whereas a genotype-first analysis can provide a lower limit. Recent studies of monogenic cancer disorders in the clinically unselected cohort also observed reduced penetrance^29^. It has been suggested that the reduced penetrance seen in clinically unselected cancer cases was due to genetic heterogeneity or even inclusion of non-pathogenic variants in the analysis^30,31^. We have excluded this explanation in our analysis through robust definition of pathogenic variants and multiple sensitivity analyses including restricting the analysis to a relatively common lower penetrance variant in *HNF4A*.

The reduced penetrance of *HNF1A*/*4A*-MODY in clinically unselected cohorts was largely consistent with the lower background rate of diabetes in these cohorts. The penetrance and background rate of diabetes was highest in the family members of probands followed by the health system-based cohort (Geisinger) and lowest in the population-based cohort (UK Biobank). In line with this, the relative risk (hazard ratios) of getting diabetes was largely constant for *HNF1A*/*4A*-MODY across three cohorts suggesting that the ascertainment context largely explains the variation in penetrance seen across these cohorts. The effect of cohort ascertainment on penetrance is likely due to environmental or/and genetic modifiers^25^. We have previously shown that the presence of the common variant *HNF1A* p.(I27L) reduces the age of diagnosis of *HNF1A* MODY^32^. Recent studies of monogenic cardiomyopathy, familial hypercholesterolemia and monogenic breast cancer also observed the impact of common variants on the phenotype^33-35^. Despite the relatively large size of our cohorts, we were limited by small numbers of carriers of individual pathogenic variants and were unable to evaluate modifiers. Further studies of common variants associated with type 2 diabetes in MODY carriers from different ascertainment contexts will be useful. However, this will require even larger clinically unselected/population cohorts to increase MODY carriers for robust analysis.

In contrast, the penetrance of pathogenic variants in *GCK* is not affected by ascertainment. We suggest that this result is explained by the different pathogenic mechanism of *HNF1A/4A*-MODY versus *GCK*-MODY. Whilst *HNF1A/4A*-MODY is characterised by progressive beta-cell dysfunction leading to age-related onset of diabetes^11,36^, *GCK*-MODY, causes life-long stable mild fasting hyperglycaemia with only a modest increase with age^26,27^. The glucose in these individuals is physiologically regulated, albeit at slightly higher glucose level, and shows minor variation around this higher set point within an individual over the lifetime. Because of this, pharmacological treatment has no impact on the glucose level, and identifying an individual at a given age with high glucose simply reflects the age of assessment rather than the true onset of high glucose, which is present from birth^28^. Our data showing that the penetrance and the level of HbA1c and fasting blood glucose were largely similar across all cohorts further support this observation and also explains the lack of effect of ascertainment with *GCK*-MODY. We did observe modestly lower HbA1c in probands compared to other cohorts (2 mmol/mol), which is likely due to the young age of the probands (13y vs. >35y). We posit that *GCK*-MODY may be representative of other highly penetrant monogenic disorders with congenital onset, where ascertainment context will not be important.

Our results have important and far-reaching clinical implications in the era of the genome-wide genotype-first approach to genetic testing. The widespread use of genome/exome sequencing for clinical and research has led to incidental identification of pathogenic variants for rare monogenic disorders in individuals without relevant phenotypes or a family history of the disease. Although specific policies vary between healthcare systems, these are increasingly being reported back to patients^37,38^. Our study results are particularly relevant now that *HNF1A* is included in the “secondary finding” gene list published by the American College of Medical Genetics and Genomics (ACMG) in 2021^39^. Our results show that pathogenic *HNF1A* variants found incidentally have a substantially reduced penetrance, and fewer than half of these individuals develop diabetes by age 50. These results call into question the reporting of secondary findings from genes such as *HNF1A*, where age-dependent penetrance is highly variable and depends on ascertainment context.

Our results for *HNF1A* and *HNF4A* clearly demonstrate that the interpretation of a variant particularly causing age-related monogenic diseases must change based on the context in which it is found. If an individual is referred for MODY testing, then their diabetes is likely explained by a pathogenic variant in one of these genes. However, if the same pathogenic variant is identified from routine sequencing in a hospital setting where the patient has been referred for other reasons, or through sequencing of healthy individuals in population research studies or direct-to-consumer testing, the penetrance of diabetes is substantially lower. This finding suggests that we need to tailor genetic interpretation and counselling for individuals based on the context in which a pathogenic variant was identified. It has previously been shown that an individual with a family history of breast cancer and a pathogenic variant in *BRCA1/2* is at substantially higher risk than a *BRCA1/2* variant carrier without a family history^29,40^. In such cases, individuals with a pathogenic variant identified incidentally may not need the same level of intensive treatment and/or management as individuals identified through clinically-directed testing.

The classical definition of penetrance may need to be reimagined in light of our study and other recent studies of rare pathogenic variants outside the clinical context. Penetrance is usually defined as the absolute risk of developing a disease in an individual with a pathogenic variant, i.e., Probability (Disease|Genotype) or P(D|G)^25^. But it has classically been estimated almost exclusively from family studies of individuals with monogenic disease, i.e., P(G|D). Bayes theorem tells us that these two measures are not equal, but are related through the disease prevalence P(D) and the allele frequency P(G) *in the population being tested*. We show that even the rarest, most pathogenic variants can confer a very low absolute risk when identified through a genotype-first approach (i.e., low prior risk of disease context) whereas the same variants can have very high absolute risk when identified through a phenotype-first approach (i.e., high prior risk of disease context). However, the relative risk of the disease in individuals carrying a pathogenic variant may be similar, indicating that the biological impact of the variant is equivalent in both contexts. This result clearly highlights the limitation of the current definition of penetrance, which treats each variant out of context and does not consider the prior risk of disease. It also highlights that caution should be exercised when using a population cohort to assess the pathogenicity of a variant or gene, where lack of association may simply reflect ascertainment context rather than true biological effect.

In conclusion, using monogenic diabetes as an example, we show that penetrance of age-related onset of monogenic disorders depends on the background rate of the disease. Importantly, ascertainment context should be taken into consideration for interpretation and clinical management of individuals with a pathogenic variant that causes a monogenic disorder.

## Data Availability

All data for this study are included in the main text of the manuscript or the supplementary appendix.

## Acknowledgements

The authors would like to acknowledge the participants of the MyCode® Community Health Initiative for use of their genomic and electronic health information, without them this study would not be possible. The patient enrollment and exome sequencing for the DiscovEHR study were funded by the Regeneron Genetics Center. We thank the Geisinger-Regeneron DiscovEHR Collaboration for making the genotype data and phenotype available for this project. This research has been conducted using the UK Biobank Resource. This work was carried out under UK Biobank project number 49847.

