## Supplementary appendix for "The penetrance of age-related monogenic disease depends on ascertainment context"

Uyenlinh L Mirshahi, Ph.D.[1], Kevin Colclough, DClin.Sci.[2], Caroline F Wright, Ph.D.[3], Andrew R Wood, Ph.D.[3], Robin N Beaumont, Ph.D.[3], Jessica Tyrrell, Ph.D.[3], Thomas W Laver, Ph.D.[3], Richard Stahl, B.S.[1], Alicia Golden, B.S.[1], Jessica M Goehringer, M.S., C.G.C. [1], Geisinger-Regeneron DiscovEHR Collaboration, Timothy F Frayling, Ph.D.[3], Andrew T Hattersley, D.M.[3], David J Carey, Ph.D.[1]*, Michael N Weedon, Ph.D.[3]*, Kashyap A Patel, Ph.D.[3]*

1. Geisinger Clinic, Geisinger Health System, Danville, PA, USA
2. Molecular Genetics, Royal Devon and Exeter NHS Foundation Trust, Exeter, U.K.
3. Institute of Biomedical and Clinical Science, College of Medicine and Health, University of Exeter, Exeter, U.K.

*Equal contribution

**Regeneron Genetics Center Banner Author List and Contribution Statements**

All authors/contributors are listed in alphabetical order.

**RGC Management and Leadership Team**
Goncalo Abecasis, Aris Baras, Michael Cantor, Giovanni Coppola, Aris Economides, Luca A. Lotta, John D. Overton, Jeffrey G. Reid, Alan Shuldiner, Katia Karalis and Katherine Siminovitch

Contribution: All authors contributed to securing funding, study design and oversight. All authors reviewed the final version of the manuscript.

**Sequencing and Lab Operations**
Christina Beechert, Caitlin Forsythe, M.S., Erin D. Fuller, Zhenhua Gu, M.S., Michael Lattari, Alexander Lopez, M.S., John D. Overton, , Thomas D. Schleicher, M.S., Maria Sotiropoulos Padilla, M.S., Louis Widom, Sarah E. Wolf, M.S., Manasi Pradhan, M.S., Kia Manoochehri, Ricardo H. Ulloa.

Contribution: C.B., C.F., A.L., and J.D.O. performed and are responsible for sample genotyping. C.B, C.F., E.D.F., M.L., M.S.P., L.W., S.E.W., A.L., and J.D.O. performed and are responsible for exome sequencing. T.D.S., Z.G., A.L., and J.D.O. conceived and are responsible for laboratory automation. M.S.P., K.M., R.U., and J.D.O are responsible for sample tracking and the library information management system.

**Genome Informatics**
Xiaodong Bai, , Suganthi Balasubramanian, , Andrew Blumenfeld, Boris Boutkov, , Gisu Eom, Lukas Habegger, , Alicia Hawes, B.S., Shareef Khalid, Olga Krasheninina, M.S., Rouel Lanche, Adam J. Mansfield, B.A., Evan K. Maxwell, Mrunali Nafde, Sean O'Keeffe, M.S., Max Orelus, Razvan Panea, , Tommy Polanco, B.A., Ayesha Rasool, M.S., Jeffrey G. Reid, , William Salerno, , Jeffrey C. Staples,

Contribution: X.B., A.H., O.K., A.M., S.O., R.P., T.P., A.R., W.S. and J.G.R. performed and are responsible for the compute logistics, analysis and infrastructure needed to produce exome and genotype data. G.E., M.O., M.N. and J.G.R. provided compute infrastructure development and operational support. S.B., S.K., and J.G.R. provide variant and gene annotations and their functional interpretation of variants. E.M., J.S., R.L., B.B., A.B., L.H., J.G.R. conceived and are responsible for creating, developing, and deploying analysis platforms and computational methods for analyzing genomic data.

**Clinical Informatics:**
Michael Cantor, Dadong Li and Deepika Sharma

Contribution: All authors contributed to the clinical informatics of the project

**Research Program Management**
Marcus B. Jones, Jason Mighty, and Lyndon J. Mitnaul

Contribution: All authors contributed to the management and coordination of all research activities, planning and execution. All authors contributed to the review process for the final version of the manuscript.

**Table S1: Characteristics of the study cohorts at recruitment.** Values are mean (SD) for continuous variables or number of individuals (%) for categorical variables. The number of individuals with available data is also indicated where appropriate.

| Characteristics | MODY probands (index cases) | MODY family members | Geisinger cohort | UK Biobank |
|---|---|---|---|---|
| **N** | 1,742 | 2,194 | 132,194 | 198,748 |
| **Age, y** | 27.3 (14.6) | 35.1 (20.3) | 52.9 (17.5) | 56.9 (8.1) |
| **Female Sex, n (%)** | 1,141 (66) | 1,291 (59) | 80,956 (61) | 109,387 (55) |
| **BMI (kg/m$^2$)** | 23.9 (4.1), n=1,360 | 25.2 (4.4), n=1,016 | 31.4 (8.2), n=129,529 | 27.4 (4.7), n=197,798 |
| **Diabetes*, n (%)** | 803 (100) | 1,168 (53) | 31,266 (24) | 11,488 (6) |
| **Age at diabetes diagnosis, y** | 19.5 (9.4) | 28.0 (15.3) | 52.6(14.4) | 52.5 (12.0) |
| **Parent with diabetes, n (%)** | 1,393 (80) | 1,306 (60) | 40,393 (31) | 34,270 (17) |
| **HbA1c, mmol/mol** | 52.5 (15.7), n=1,439 | 49.8 (17.1), n=984 | 44.9 (15.3), n=58,920 | 38.2 (6.2), n=188,924 |
| **Fasting glucose, mmol/l** | 7.1 (1.9), n=958 | 6.5 (2.1), n=655 | 6.1 (2.2), n=85,437 | 5.1 (1.0), n=41,898 |
| **European ancestry, n (%)** | 1,437 (90) | 1,776 (94) | 125,850 (95) | 182,920 (92) |
| **Unrelated, n (%)** | 1,742 (100) | 814 (37) | 87,234 (66) | 184,142 (93) |

Abbreviations MODY, matured onset diabetes of the young; y, years; BMI, body mass index, *excluding GCK-MODY probands

**Table S2: Characteristics of all family members at recruitment by each gene.** Values are mean (SD) for continuous variables or number of individuals (%) for categorical variables. The number of individuals with available data is also indicated where appropriate.

| Characteristics | ***HNF1A*** | ***HNF4A*** | ***GCK*** |
|---|---|---|---|
| **N** | 954 | 253 | 987 |
| **Age at recruitment, y** | 36.6 (19.1) | 24.3 (20.9) | 33.9 (21.3) |
| **Female Sex, n (%)** | 562 (59) | 154 (61) | 575 (58.3) |
| **BMI at recruitment (kg/m$^2$)** | 25.2 (4.2), n=536 | 24.3 (20.9), n=253 | 25.1 (4.7), n=370 |
| **Diabetes, n (%)** | 586 (61) | 125 (49) | 457 (46.3) |
| **Age at diabetes diagnosis, y** | 25.7 (13.8) | 27.1 (12.6) | 32.1 (17.4) |
| **Parent with diabetes, n (%)** | 681 (71) | 182 (72) | 443 (44.9) |
| **HbA1c, mmol/mol** | 50.9 (20.4), n=486 | 55.6 (17.4), n=89 | 47.1 (11.1), n=409 |
| **Random Glucose, mmol/l** | 16.8 (8.7), n=7 | 8.3 (7.2), n=6 | 9.4 (6.9), n=12 |
| **Fasting glucose, mmol/l** | 6.7 (3.1), n=199 | 5.9 (2.6), n=34 | 6.5 (1.3), n=422 |
| **European Ancestry, n (%)** | 840 (95), n=840 | 190 (90), n=211 | 746 (93.4), n=799 |
| **Unrelated, n (%)** | 328 (34) | 86 (34) | 400 (40.5) |

**Table S3. Pathogenic variants of *HNF1A, HNF4A*, and *GCK* in MODY proband, proband family members, Geisinger cohort, and UK Biobank.** This table is included in Excel format.

**Table S4: Characteristics of *HNF1A* carriers in MODY probands, proband family members, Geisinger cohort, and UK Biobank at recruitment.** Values are mean (SD) for continuous variables or number of individuals (%) for categorical variables. The number of individuals with available data is also indicated where appropriate.

| Characteristics | MODY probands (index cases) | MODY family members | Geisinger cohort | UK Biobank |
|---|---|---|---|---|
| **N** | 661 | 622 | 13 | 22 |
| **Age, y** | 31.7 (15.0 | 36.8 (18.3) | 47.8 (20.1) | 56.1 (8.5) |
| **Female Sex, n (%)** | 455 (69) | 366 (59) | 10 (71) | 14 (64) |
| **BMI (kg/m$^2$)** | 24.6 (4.0), n=527 | 24.8 (3.9), n=367 | 27.3 (6.9) | 26.2 (4.1) |
| **Diabetes, n (%)** | 661 (100) | 526 (85) | 9 (64) | 11 (50) |
| **Age at diabetes diagnosis, y** | 19.3 (7.8) | 23.9 (11.8) | 36.6 (15.6) | 32.0 (16.8) |
| **Parent with diabetes, n (%)** | 582 (88) | 491 (79) | 8 (57) | 14 (64) |
| **HbA1c, mmol/mol** | 59.2 (19.0), n=553 | 56.5 (19.1), n=355 | 56.3 (12.4), n=10 | 46.4 (12.7), n=21 |
| **Fasting glucose, mmol/l** | 8.3 (3.1), n=193 | 7.7 (3.6), n=119 | 7.7 (2.7) | 5.1 (1.1), n=5 |
| **European ancestry, n (%)** | 564 (90) | 546 (93) | 14 (100) | 22 (100) |
| **Unrelated, n (%)** | 661 (100) | 250 (40) | 12 (86) | 21 (95) |
| **PTV, n (%)** | 374 (57) | 350 (56) | 9 (64) | 8 (36) |
| **Missense, n (%)** | 287 (43) | 272 (44) | 5(36) | 14 (64) |

Abbreviations MODY, matured onset diabetes of the young; y, years; BMI, body mass index; PTV, putative truncating variants

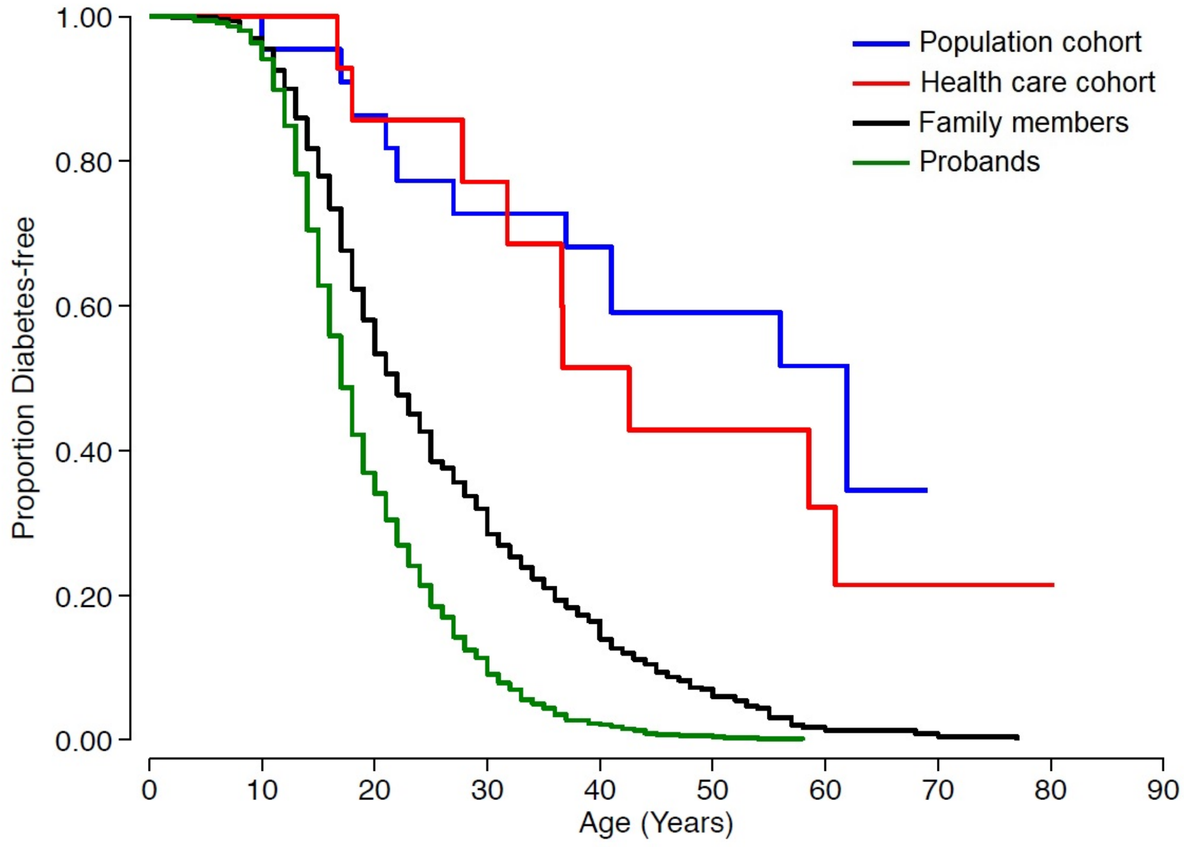

**Figure S1: Penetrance of diabetes for pathogenic *HNF1A* variant carriers in clinically selected and unselected cohorts.** Kaplan-Meier survival curves of diabetes for *HNF1A*-MODY probands (N=661), their family members with pathogenic *HNF1A* variants (N=622), and carriers of pathogenic *HNF1A* variants from health care-based Geisinger cohort (N=13) and UK Biobank population cohort (N=22). The log rank test p values for probands versus each unselected cohort were $3x10^{-26}$, $3x10^{-09}$, $5x10^{-16}$, respectively.

**Table S5: Univariate and multivariate Cox proportional hazard ratios for diabetes for *HNF1A* carriers in each unselected cohort relative to *HNF1A*-MODY probands.** Hazard ratios (95% CI), p values, and number of all carriers (top) and unrelated carriers of European ancestry (bottom) in each cohort in univariate (left) and multivariate (right) regression analysis vs. probands (base). Multivariate regression adjusted for age at recruitment, sex, body mass index, family history of diabetes, and variant type.

| | **Unadjusted** | | | **Adjusted*** | | |
|---|---|---|---|---|---|---|
| | **Hazard Ratio (95% CI)** | **P Value** | **N** | **Hazard Ratio (95% CI)** | **P Value** | **N** |
| **All Carriers** | | | | | | |
| **Probands** | Base | | 661 | Base | | 527 |
| **Family members of proband** | 0.6 (0.5 - 0.6) | 1.1E-23 | 622 | 0.7 (0.6 - 0.9) | 5.7E-05 | 367 |
| **Geisinger cohort** | 0.2 (0.1 - 0.3) | 7.7E-07 | 14 | 0.2 (0.1 - 0.4) | 6.2E-05 | 14 |
| **UK Biobank** | 0.1 (0.1 - 0.2) | 2.8E-11 | 22 | 0.2 (0.1 - 0.3) | 5.5E-07 | 22 |
| **Unrelated European ancestry carriers** | | | | | | |
| **Probands** | Base | | 564 | Base | | 456 |
| **Unrelated Family members of proband** | 0.5 (0.4 - 0.6) | 3.8E-17 | 219 | 0.8 (0.6 - 1.0) | 0.04 | 134 |
| **Unrelated Geisinger cohort** | 0.1 (0.1 - 0.3) | 1.7E-06 | 12 | 0.2 (0.1 - 0.4) | 9.8E-05 | 12 |
| **Unrelated UK Biobank** | 0.1 (0.1 - 0.2) | 2.0E-10 | 21 | 0.2 (0.1 - 0.4) | 1.8E-06 | 21 |

*Adjusted for age at study, sex, BMI, parent with diabetes and variant type (PTV vs. missense)
Abbreviations CI, confidence intervals

**Table S6: Characteristics of *HNF4A* carriers in MODY probands, proband family members, Geisinger cohort and UK Biobank at recruitment.** Values are mean (SD) for continuous variables or number of individuals (%) for categorical variables. The number of individuals with available data is also indicated where appropriate.

| Characteristics | MODY probands (index cases) | MODY family members | Geisinger cohort | UK Biobank |
|---|---|---|---|---|
| **N** | 142 | 169 | 20 | 29 |
| **Age, y** | 33.1 (14.6) | 35.8 (21.0) | 48.6 (14.7) | 56.8 (7.1) |
| **Female Sex, n (%)** | 103 (73) | 101 (60) | 7 (35) | 18 (62) |
| **BMI (kg/m2)** | 25.5 (4.6), n=127 | 25.7 (4.9), n=89 | 31.1 (5.1) | 26.8 (4.4), n=28 |
| **Diabetes, n (%)** | 142 (100) | 114 (67) | 9 (45) | 11 (38) |
| **Age at diabetes diagnosis, y** | 21.6 (8.3) | 26.6 (12.0) | 48.1 (12.0) | 43.5 (14.7) |
| **Parent with diabetes, n (%)** | 122 (86) | 139 (82) | 5 (25) | 13 (45) |
| **HbA1c, mmol/mol** | 62.4 (22.2), n=118 | 57.4 (16.9), n=74 | 50.7 (14.1), n=13 | 45.3 (12.4), n=27 |
| **Fasting glucose, mmol/l** | 9.5 (3.8), n=38 | 6.4 (3.0), n=23 | 7.7 (4.4), n=17 | 4.8 (0.7), n=7 |
| **European ancestry, n (%)** | 122 (89) | 131 (90) | 20 (100) | 26 (90) |
| **Unrelated, n (%)** | 142 (100) | 66 (39) | 18 (90) | 27 (93) |
| **PTV, n (%)** | 35 (25) | 43 (25) | 14 (70) | 4 (14) |
| **Missense, n (%)** | 107 (75) | 126 (75) | 6 (30) | 25 (86) |

Abbreviations MODY, matured onset diabetes of the young; y, years; BMI, body mass index; PTV, putative truncating variants

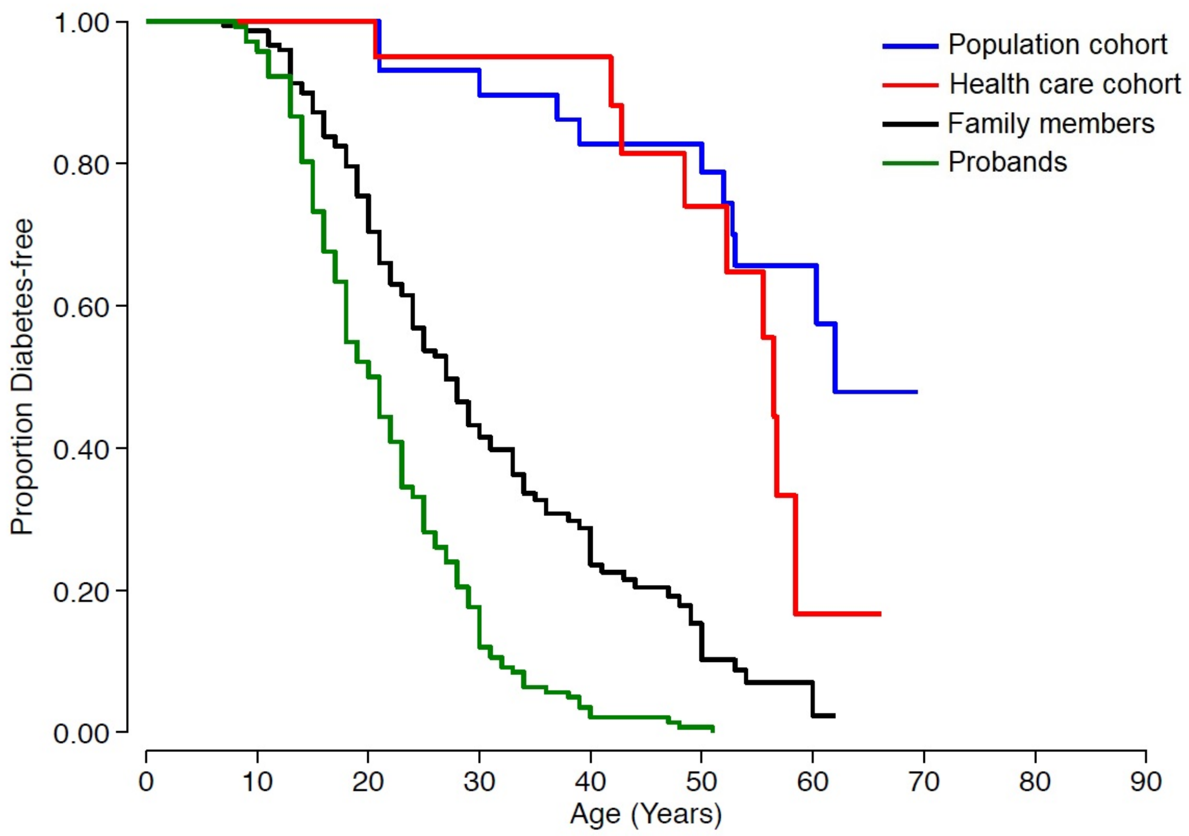

**Figure S2. Penetrance of diabetes for pathogenic *HNF4A* variant carriers in clinically selected and unselected cohorts.** Kaplan-Meier survival curves for diabetes for *HNF4A*-MODY probands (N=142, their family members with pathogenic *HNF4A* variants (N=169), and carriers of pathogenic *HNF4A* variants from health care-based Geisinger cohort (N=20) and UK Biobank population cohort (N=29). The log rank test p values probands versus each unselected cohort were $8 \times 10^{-11}$, $2 \times 10^{-14}$, $3 \times 10^{-19}$, respectively.

**Table S7: Univariate and multivariate Cox proportional regression hazard ratios for diabetes for *HNF4A* carriers in each unselected cohort relative to *HNF4A*-MODY probands.** Hazard ratios (95% CI), p values, and number of all carriers (top) and unrelated carriers of European ancestry (bottom) in each cohort in univariate (left) and multivariate (right) regression analysis vs. probands (base). Multivariate regression adjusted for age at recruitment, sex, body mass index, family history of diabetes, and variant type.

| | Unadjusted | | | Adjusted* | | |
|---|---|---|---|---|---|---|
| | Hazard Ratio (95% CI) | P Value | N | Adjusted Hazard Ratio (95% CI)* | Adjusted P Value* | N |
| **All Carriers** | | | | | | |
| **Probands** | Base | | 142 | Base | | 127 |
| **Family members of proband** | 0.4 (0.3 - 0.6) | 8.1E-10 | 169 | 0.7 (0.5 - 1.0) | 0.03 | 89 |
| **Geisinger cohort** | 0.05 (0.02-0.13 | 8.2E-09 | 20 | 0.05 (0.02 - 0.2) | 5.5E-07 | 20 |
| **UK Biobank** | 0.05 (0.02 - 0.1) | 6.9E-12 | 29 | 0.08 (0.02 - 0.2) | 1.7E-06 | 28 |
| **Unrelated European ancestry carriers** | | | | | | |
| **Probands** | Base | | 122 | Base | | 110 |
| **Unrelated Family members of proband** | 0.5 (0.4 - 0.7) | 1.0E-04 | 56 | 1.3 (0.8 - 2.0) | 0.3 | 36 |
| **Unrelated Geisinger cohort** | 0.05(0.02 - 0.2) | 3.8E-08 | 18 | 0.06(0.02 - 0.2) | 5.1E-08 | 18 |
| **Unrelated UK Biobank** | 0.06(0.02 - 0.1) | 1.7E-10 | 25 | 0.09 (0.03 - 0.2) | 6.2E-06 | 24 |

*Adjusted for age at study, sex, body mass index, parent with diabetes and variant type (PTV vs. missense)

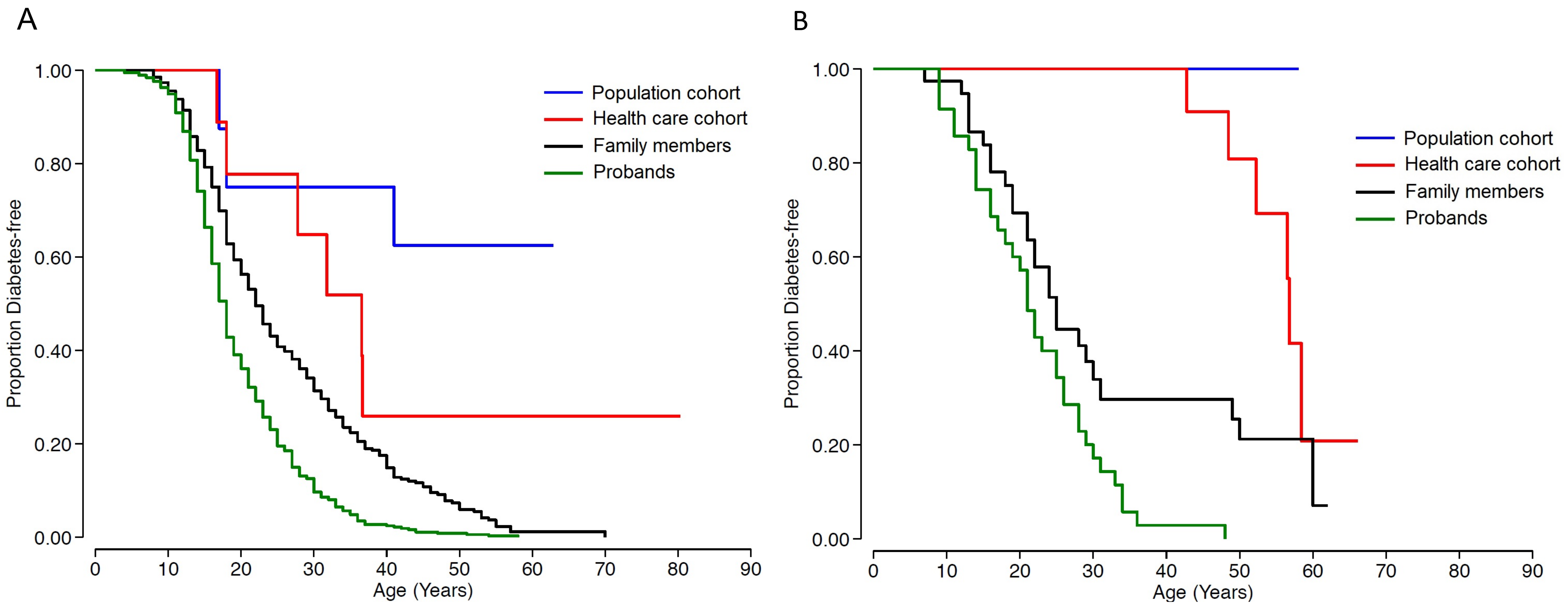

**Figure S3. Penetrance of diabetes for *HNF1A* and *HNF4A* putative truncating variants (PTVs) observed in both carriers from clinically selected and unselected cohorts.** Kaplan-Meier survival curves of diabetes for A) *HNF1A*-MODY in probands (N=374), their family members (N=350), healthcare-based Geisinger cohort (N=9), and UK Biobank population cohort (N=8) restricted to PTVs seen in all those cohorts only. The log rank test p values verses probands for each unselected cohort were $2x10^{-14}$, $1x10^{-04}$, $2x10^{-07}$, respectively. B) Same as A except for *HNF4A*-MODY; probands (N=35), their family members with *HNF4A* PTVs (N=43), and *HNF4A* PTV carriers from healthcare-based Geisinger cohort (N=14) and UK Biobank population cohort (N=4). The log rank test p values verses probands for each unselected cohort were 0.01, $2x10^{-09}$, 0.0002, respectively.

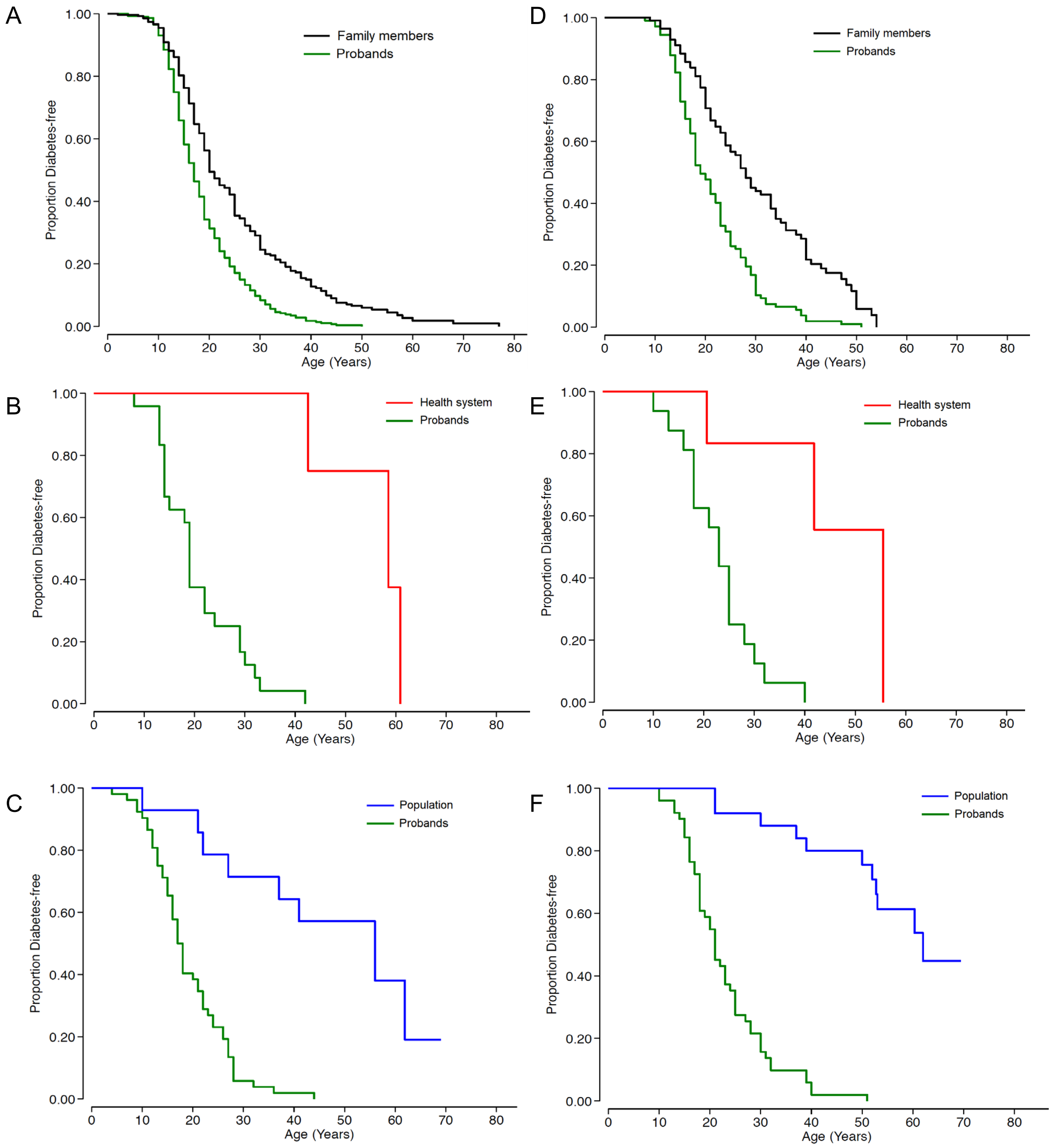

**Figure S4. Penetrance of diabetes missense *HNF1A*-MODY and *HNF4A*-MODY restricted to variants observed in carriers from unselected comparison cohort and probands.** Left: Kaplan-Meier survival curves of diabetes for *HNF1A*-MODY for A) proband family members (N=272) vs. probands (N=287), B) Geisinger healthcare system (N=5) vs. probands (N=287), and C) UK Biobank population (N=14) vs. probands (N=287). The log rank test p values verses probands for each unselected cohort were 3.2x10$^{-12}$, 2.3x10$^{-04}$, 5.9x10$^{-07}$, respectively. Right: Kaplan Meier survival curves of diabetes for *HNF4A*-MODY for A) proband family members (N=126) vs. probands (N=107), B) Geisinger (N=6) vs. probands (N=16), and C) UK Biobank (N=25) vs. probands (N=51). The log rank test p values verses probands for each unselected cohort were 3.9x10$^{-09}$, 1.6x10$^{-03}$, 7.4x10$^{-13}$, respectively.

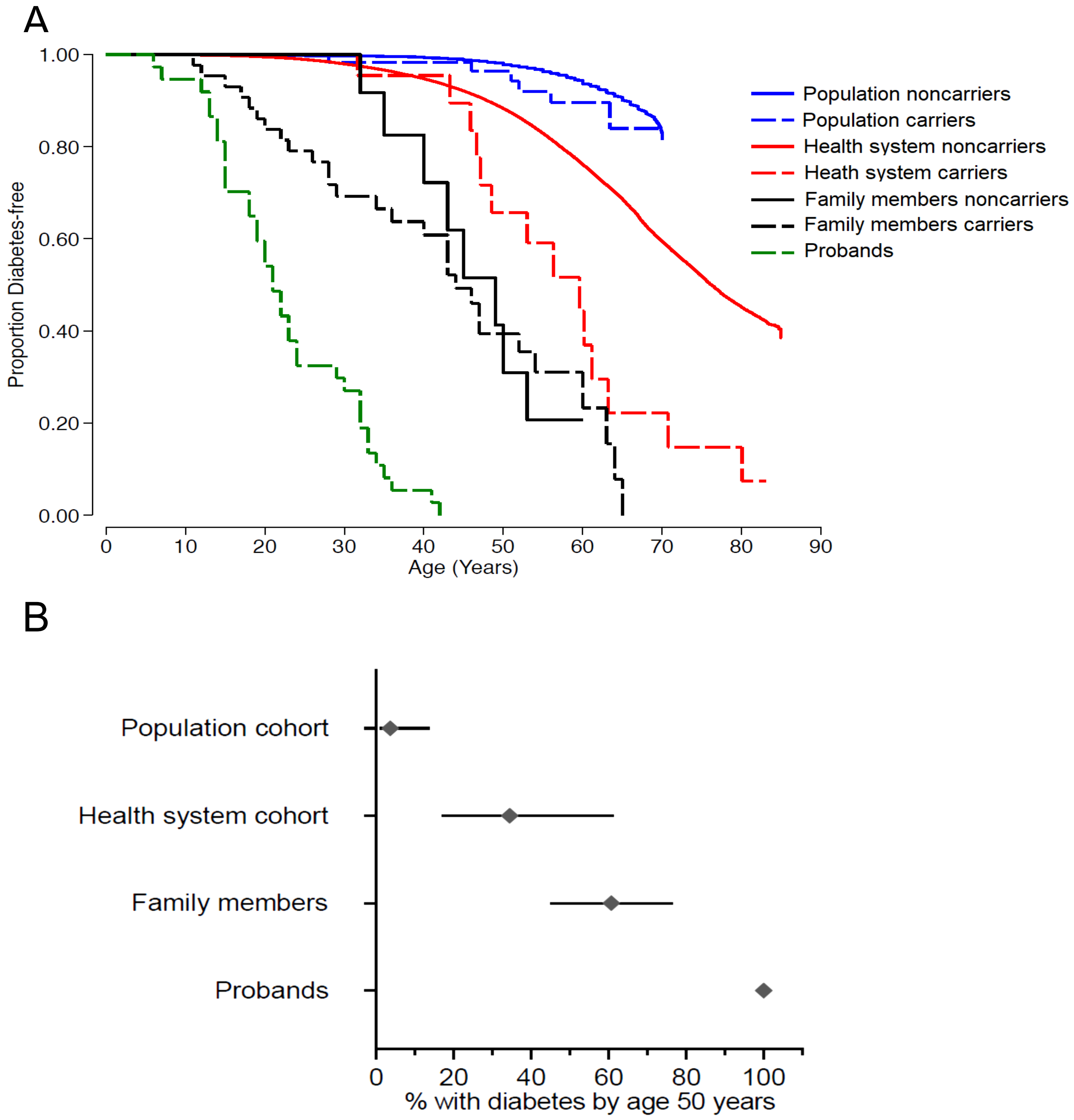

**Figure S5. Penetrance of diabetes for pathogenic *HNF4A* p.R114W variant carriers and noncarriers in clinically selected and unselected cohorts.** A) Kaplan Meier survival curves of diabetes for *HNF4A*-MODY p.R114W pathogenic variant probands (N=37), their family member carriers (N=43) and noncarriers (N=41) of *HNF4A* R114W, carriers (N=24) and noncarriers (N=132,150) of *HNF4A* R114W from Geisinger healthcare system cohort, and carriers (N=58) and noncarriers (N=198,661) of *HNF4A* R114W from UK Biobank population cohort. The log rank test p value for probands versus carriers of proband family members was $1x10^{-09}$, carriers of Geisinger cohort was $1x10^{-13}$, and carriers of UK Biobank $5x10^{-29}$. Results were similar when analysed with unrelated individuals of European ancestry with and without adjustment for age at study, sex, BMI, parents with diabetes status, and variant types (see Supplemental table 8). B) Penetrance of diabetes for pathogenic *HNF4A* p.R114W variant carriers in all four cohorts at age 50 years with 95% CI.

**Table S8: Univariate and multivariate Cox proportional regression hazard ratios for diabetes for *HNF4A* p.(R114W) carriers in each unselected cohort relative to *HNF4A* p.(R114W) MODY probands.** Hazard ratios (95% CI), p values, and number of all carriers (top) and unrelated carriers of European ancestry (bottom) in each cohort in univariate (left) and multivariate (right) regression analysis vs. probands (base). Multivariate regression adjusted for age at recruitment, sex, body mass index, family history of diabetes, and variant type.

| | Hazard Ratio (95% CI) | P Value | N | Adjusted Hazard Ratio (95% CI)* | Adjusted P Value* | N |
|---|---|---|---|---|---|---|
| **All Carriers** | | | | | | |
| **Probands** | Base | | 37 | Base | | 31 |
| **Family members of proband** | 0.2 (0.1 - 0.3) | 5.4E-08 | 43 | 0.4 (0.2 - 0.9) | 0.02 | 24 |
| **Geisinger cohort** | 0.01 (0.002 - 0.1) | 2.4E-05 | 24 | 0.02 (0.002 - 0.2) | 5.9E-04 | 24 |
| **UK Biobank** | 0.004 (0.0006 - 0.04) | 2.2E-07 | 58 | 0.009 (-0.0008 - 0.09) | 6.4E-05 | 28 |
| **Unrelated European ancestry carriers** | | | | | | |
| **Probands** | Base | | 33 | Base | | 29 |
| **Unrelated Family members of proband** | 0.3 (0.1 - 0.6) | 0.003 | 15 | 0.7 (0.3 - 2.0) | 0.6 | 11 |
| **Unrelated Geisinger cohort** | 0.02 (0.002 - 0.1) | 8.8E-05 | 19 | 0.03 (0.003 - 0.3) | 2.5E-03 | 19 |
| **Unrelated UK Biobank** | 0.005 (0.0007 - 0.04) | 3.9E-07 | 54 | 0.009 (0.0009 - 0.1) | 7.8E-05 | 53 |

*Adjusted for age at study, sex, body mass index, parent with diabetes and variant type (PTV vs. missense)

**Table S9: Cox proportional hazard ratios for age related onset of diabetes for *HNF1A* or *HNF4A* carriers relative to non-carriers in each clinically unselected study cohort.** Relative hazard ratios (95% CI) for diagnosis of diabetes in carriers of *HNF1A*, *HNF4A*, or *HNF4A* R114W vs. noncarriers in each cohort. Cox regression models were applied to all individuals in the cohort or unrelated individuals of European ancestry. Meta-analysis using random-effects models by each condition showed no statistically significant differences in relative hazard ratios for diabetes between cohorts with *HNF4A* but a significant difference with *HNF1A*, although the relative hazard ratios 95% CI between cohorts overlap.

| | Hazard Ratio (95% CI) | *P* for Heterogeneity |
|---|---|---|
| **All HNF1A** | | |
| **Family members** | 11.0 (8.3 - 14.7) | |
| **Geisinger cohort** | 4.0 (2.1 - 7.7) | 0.005 |
| **UK Biobank** | 15.6 (8.7 - 28.2) | |
| **Unrelated European HNF1A** | | |
| **Family members** | 7.8 (4.9 - 12.5) | |
| **Geisinger cohort** | 3.8 (1.9 - 7.6) | 0.002 |
| **UK Biobank** | 18.6 (10.3 - 33.7) | |
| **All HNF4A** | | |
| **Family members** | 7.6 (4.1 - 14.3) | |
| **Geisinger cohort** | 3.5 (1.8 - 6.7) | 0.15 |
| **UK Biobank** | 7.7 (4.3 - 13.9) | |
| **Unrelated European HNF4A** | | |
| **Family members** | 2.8 (1.2 - 6.3) | |
| **Geisinger cohort** | 4.0 (2.1 - 7.7) | 0.08 |
| **UK Biobank** | 8.7 (4.5 - 17) | |
| **All HNF4A p.R114W** | | |
| **Family members** | 1.3 (0.6 - 2.8) | |
| **Geisinger cohort** | 2.9 (1.7 - 4.9) | 0.2 |
| **UK Biobank** | 1.7 (0.8 - 3.7) | |
| **Unrelated European HNF4A p.R114W** | | |
| **Family members** | 2.2 (0.6 - 7.8) | |
| **Geisinger cohort** | 4.3 (2.5 - 7.4) | 0.2 |
| **UK Biobank** | 2.0 (0.9 - 4.5) | |

**Table S10: Characteristics of *GCK* carriers in MODY probands, proband family members, Geisinger cohort and UK Biobank at recruitment.** Values are mean (SD) for continuous variables or number of individuals (%) for categorical variables. The number of individuals with available data is also indicated where appropriate.

| Characteristics | MODY probands (index cases) | MODY family members | Geisinger cohort | UK Biobank |
|---|---|---|---|---|
| **N** | 939 | 723 | 32 | 83 |
| **Age, y** | 13.2 (13.2) | 35.7 (20.7) | 55.4 (18.9) | 56.9 (7.9) |
| **Female Sex, n (%)** | 583 (62) | 432 (60) | 18 (56) | 49 (59) |
| **BMI ($kg/m^2$)** | 3.9 (3.9), n=706 | 24.7 (4.5), n=290 | 30.5 (7.4), n=31 | 27.4 (4.7) |
| **Diabetes, n (%)** | 602 (64) | 418 (58) | 22 (69) | 54 (65) |
| **Age at diabetes diagnosis, y** | 689 (73) | 362 (50) | 12 (38) | 34 (41) |
| **Parent with diabetes, n (%)** | 6.1 (6.1), n=768 | 48.4 (8.4), n=331 | 48.3 (9.9), n=26 | 47.5 (4.6), n=83 |
| **HbA1c, mmol/mol** | 0.8 (0.8), n=727 | 6.9 (1.1), n=323 | 6.9 (1.3), n=22 | 6.5 (1.1), n=17 |
| **Fasting glucose, mmol/l** | 751 (90) | 562 (93) | 32 (100) | 80 (96) |
| **European ancestry, n (%)** | 939 (100) | 330 (46) | 27 (84) | 75 (90) |
| **Unrelated, n (%)** | 220 (23) | 174 (24) | 6 (19) | 21 (25) |
| **PTV, n (%)** | 719 (77) | 549 (76) | 26 (81) | 62 (75) |

**Table S11: Comparison of HbA1c (mmol/mol) in *GCK*-MODY probands vs *GCK* carriers in each unselected cohort.** Mean (95% CI) and number of carriers (N) in all carriers (top) and unrelated carriers of European ancestry (bottom) are listed. Univariate, age-adjusted, and multivariate Cox-proportional regression analyses compared HbA1c levels between each cohort and probands (base). The mean difference of HbA1c (95% CI) and p values between each cohort compared to probands is shown.

| | | | Unadjusted | | Adjusted for age | | Adjusted for multi-variables* | |
|---|---|---|---|---|---|---|---|---|
| | N | Mean (95%CI) | Mean difference vs proband (95% CI) | P vs proband | Mean difference vs proband (95% CI) | P vs proband | Mean difference vs proband (95% CI) | P vs proband |
| **All carriers** | | | | | | | | |
| **Proband** | 768 | 46.1 (45.7 - 46.6) | base | base | base | base | base | base |
| **Family members** | 331 | 48.4 (47.5 - 49.3) | 2.3 (1.4, 3.2) | 4.3E-07 | 1.5 (0.6, 2.4) | 0.002 | 1.1 (-0.1, 2.2) | 0.07 |
| **Geisinger cohort** | 26 | 48.3 (44.3 - 52.3) | 2.1 (-0.6, 4.8) | 0.1 | 0.1 (-2.7, 2.9) | 1.0 | -0.5 (-3.5, 2.5) | 0.7 |
| **UK Biobank** | 83 | 47.5 (46.5 - 48.5) | 1.3 (-0.2, 2.9) | 0.1 | -0.6 (-2.4, 1.1) | 0.5 | -0.9 (-2.8, 1.0) | 0.3 |
| **European unrelated** | | | | | | | | |
| **Proband** | 630 | 46.1 (45.6 - 46.6) | base | base | base | base | base | base |
| **Family members** | 131 | 49.5 (48.2 - 50.8) | 3.4 (2.3, 4.6) | 2.0E-08 | 2.6 (1.3, 4.0) | 0.0002 | 1.1 (-0.1, 2.2) | 0.07 |
| **Geisinger cohort** | 22 | 46.6 (43.3 - 49.5) | 0.3 (-2.4, 3.0) | 0.8 | -0.9 (-3.8, 1.9) | 0.5 | -0.5 (-3.5, 2.5) | 0.7 |
| **UK Biobank** | 74 | 47.6 (46.6 - 48.7) | 1.5 (-0.02, 3.0) | 0.05 | 0.3 (-1.5, 2.1) | 0.7 | -0.9 (-2.8, 1.0) | 0.3 |

*Adjusted for age at study, sex, and body mass index

**Table S12: Comparison of fasting blood glucose (mmol/l) in *GCK*-MODY probands vs *GCK* carriers in each unselected cohort.** Mean (95% CI) and number of carriers (N) in all carriers (top) and unrelated carriers of European ancestry (bottom) are listed. Univariate, age-adjusted, and multivariate Cox-proportional regression analyses compared fasting blood glucose levels between each cohort and probands (base). The mean difference of fasting blood glucose (95% CI) and p values between each cohort compared to probands is shown.

| | | | **Unadjusted** | | **Adjusted for age** | | **Adjusted for multi-variables*** | |
|---|---|---|---|---|---|---|---|---|
| | N | Mean (95%CI) | Mean difference vs proband (95% CI) | P vs proband | Mean difference vs proband (95% CI) | P vs proband | Mean difference vs proband (95% CI) | P vs proband |
| **All carriers** | | | | | | | | |
| **Proband** | 727 | 6.7 (6.7 - 6.8) | base | base | base | base | base | base |
| **Family members** | 323 | 6.9 (6.8 – 7.0) | 0.2 (0.04,0.3) | 0.01 | 0.08 (-0.04,0.2) | 0.2 | -0.03(-0.2,0.1) | 0.7 |
| **Geisinger cohort** | 22 | 6.9 (6.3 - 7.5) | 0.2 (-0.2,0.6) | 0.3 | -0.08 (-0.5,0.3) | 0.7 | -0.5 (-0.9,-0.02) | 0.04 |
| **UK Biobank** | 17 | 6.5 (6 - 7.1) | -0.2 (-0.6,0.2) | 0.4 | -0.4 (-0.8,0.0) | 0.07 | -0.6 (-1.1,-0.2) | 0.01 |
| **European unrelated** | | | | | | | | |
| **Proband** | 601 | 6.7 (6.7 - 6.8) | base | base | base | base | base | base |
| **Family members** | 106 | 7.0 (6.8 - 7.2) | 0.3 (0.1,0.4) | 8.30E-04 | 0.2 (0.1,0.4) | 0.01 | 0.1 (-0.1,0.4) | 0.3 |
| **Geisinger cohort** | 18 | 7.0 (6.4 - 7.6) | 0.3 (-0.1,0.6) | 0.2 | 0.2 (-0.2,0.6) | 0.3 | -0.1 (-0.5,0.4) | 0.8 |
| **UK Biobank** | 15 | 6.6 (5.9 - 7.2) | -0.1 (-0.5,0.3) | 0.5 | -0.2 (-0.6,0.2) | 0.4 | -0.4 (-0.8,0.1) | 0.1 |

*Adjusted for age at study, sex, and body mass index

**Table S13: Penetrance of mild hyperglycaemia in *GCK* carriers in unrelated European.** Proportion of *GCK* carriers with hyperglycaemia as defined by HbA1c > 39 mmol/mol (5.7%) or fasting blood glucose > 5.6 mmol/L (32.7 mmol/mol). Fisher's exact test p values compared each cohort vs. probands.

| | % (95%CI) | P vs Probands |
|---|---|---|
| **Probands** | 97 (96 - 98) | base |
| **Family members** | 96 (94 - 98) | 0.7 |
| **Geisinger cohort** | 89 (71 - 98) | 0.05 |
| **UK Biobank** | 96 (90 - 99) | 0.5 |
